# Circulatory Cytokines and Chemokines Profile in Human Coronaviruses: A systematic review and meta-analysis

**DOI:** 10.1101/2021.02.16.21251673

**Authors:** Ayat Zawawi, Abdallah Y Naser, Hassan Alwafi, Faisal Minsahwi

**Author notes:** **Correspondence author:** Dr. Faisal Minshawi, Assistant Professor of Immunology, Laboratory Medicine, Faculty of Applied Medical Science, Umm Al-Qura University, Abidiyyah, Makkah, KSA.

## Abstract

**Background:** SARS, MERS, and COVID-19 share similar characteristics as the genetic homology of SARS-CoV-2 compared to SARS-CoV and MERS-CoV is 80% and 50% respectively and cause similar clinical features. Uncontrolled release of proinflammatory mediators (cytokine storm) by activated immune cells in SARS, MERS, and COVID-19 leads to severe phenotype development.

**Aim:** This systematic review and meta-analysis aimed to evaluate the inflammatory cytokines profile associated with severe human coronavirus diseases, including three stains: MERS-CoV, SARS-CoV SARS-CoV-2, in severe patients.

**Method:** PubMed, Embase, and Cochrane Library databases were searched up to July 2020. Randomized and Observational studies reporting the inflammatory cytokines associated with severe and non-severe human coronavirus diseases, including MERS-CoV, SARS-CoV SARS-CoV-2 were included. Two reviewers independently screened articles, extracted data, and assessed the quality of included studies. Meta-analysis was performed using a random-effects model with a 95% confidence interval (CI) to estimate the pooled mean of inflammatory biomarkers.

**Results:** A high level of circulating IL-6 could be associated with the severity of the three strains of coronaviruses infection. TNF, IL-10, and IL-8 is associated with the severity of COVID-19. Increased circulating levels of CXCL10/IP10 and CCL2/MCP-1 might also be related to the severity of MERS.

**Conclusion:** This study suggests that the immune response and immunopathology in the three severe human coronavirus strains are similar to some extent. These findings highlight that nearly all studies reporting severe cases of SARS, MERS, and COVID-19 have been associated with elevated levels of IL-6, which could be used as a potential therapeutic target to improve patients’ outcomes in severe cases.

## Introduction

Coronavirus disease 2019 (COVID-19), has been characterised as the third human coronavirus (hCoV) outbreak in only two decades (1). Coronaviruses are enveloped, single-stranded RNA viruses that infect the lower respiratory tract (2). Severe Acute Respiratory Syndrome (SARS) was the first global epidemic of the twenty-first century caused by SARS-CoV (3, 4). For the first time in Hong Kong, in March 2003, the disease broke out and has spread through the world, including Asia, Europe, and the United States (5). Nearly a decade following the SARS epidemic, the Middle East Respiratory Syndrome (MERS) caused by MERS-CoV emerged in June 2012, in Saudi Arabia. The MERS outbreak also rapidly spread to more than 20 countries (6). Bats have been known to be the natural reservoir of SARS and MERS. Dromedary camels have also been found to be the zoonotic reservoir to transmit MERS to humans (7). COVID-19 is the newest emerging infectious disease (8). The infection began in Wuhan, China, in December 2019, (9). The novel coronavirus has infected over 100 million people with around 2 million death around the world (10), causing a more serious global threat than either of its two predecessors.

These three human coronaviruses share similar characteristics as the genetic homology of SARS-CoV-2 compared to SARS-CoV and MERS-CoV is 80% and 50%, respectively (11). These three respiratory infectious diseases are also associated with significant morbidity and mortality as they are highly transmissible, primary through respiratory droplets and close contact (12). However, MERS is relatively inefficiently transmitted to humans compared to SARS and COVID-19 (13, 14). Conversely, SARS and COVID-19 were characterised by a relatively low mortality rate than MERS (15). This is likely related to the viral kinetic; older people are more common to get MERS infection and comorbid illness (15).

The three respiratory infectious diseases SARS, MERS, and COVID-19 cause similar clinical features ranged from asymptomatic to severe illness that requires intubation and intensive care management (16, 17). The severity of these diseases is linked to age and chronic illness, including diabetes, hypertension, cardiovascular disease, chronic respiratory disease, and cancer (18). Whereas viral pneumonia, acute respiratory distress syndrome, and multiple organ failure are common in these diseases’ later stages (19-21). Other non-specific laboratory tests including leukopenia, lymphopenia, thrombocytopenia, and elevated serum amino transaminases, are also common in SARS, MERS, and COVID-19 infections (22).

It has also been reported that uncontrolled release of proinflammatory mediators (cytokine storm) by activated immune cells in SARS (23), MERS (24) and COVID-19 (25) leads to the development of severe phenotype. Given the potential overlap in presentation and manifestation among severe hCoVs infection, as such absence of effective treatment, it is essential to understand the immunopathological aspect such as cytokine storm syndrome (CSS). Therefore, this systematic review and meta-analysis aimed to compare the inflammatory biomarkers associated with severe human coronavirus diseases, including three significant stains: MERS-CoV, SARS-CoV, SARS-CoV-2, which may indicate a distinct inflammatory profile and, accordingly, identifying potential treatment options in severe patients.

## Methods

The systematic review and meta-analysis were carried out following the Meta-analysis of Observational Studies in Epidemiology (MOOSE) guidelines (26) and reported following the Preferred Reporting Items for Systematic reviews and Meta-Analyses (PRISMA) statement (27). The protocol of the study was registered with PROSPERO (Provisional registration number CRD42020209931).

### Databases and search strategy

An extensive search strategy summarised in Figure 1 was developed to identify relevant studies. A detailed electronic search on bibliographic databases including Medline, Embase, and Cochrane Library databases was performed from inception to July 2020. Keywords, Emtree and MeSH terms were used with both English and American spellings. The search strategy covered the following keywords: cytokines, inflammatory biomarkers, SARS-CoV, SARS-CoV-2, Severe Acute Respiratory Syndrome, Middle East Respiratory Syndrome Coronavirus, and COVID-19.

**Figure 1.**
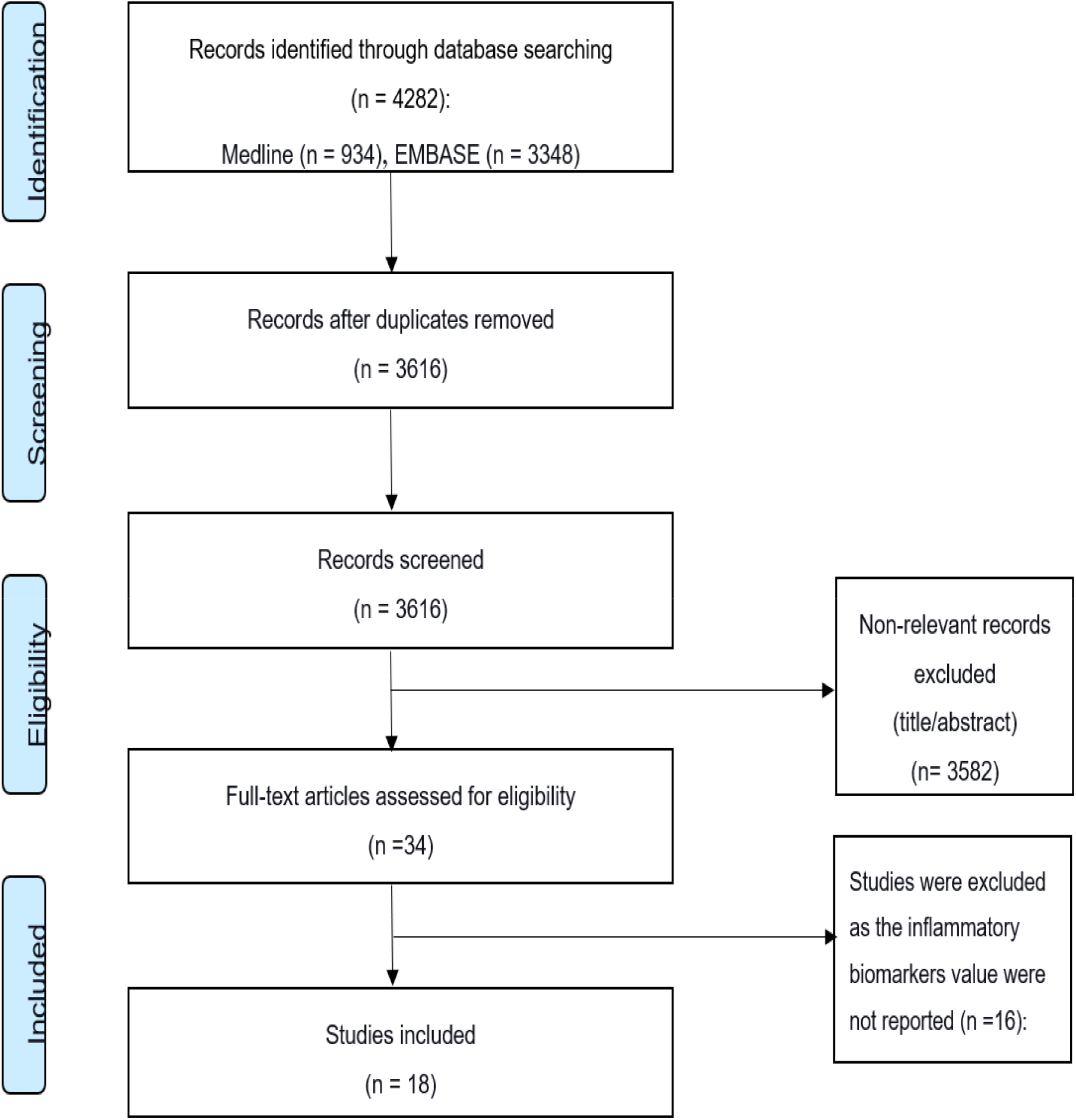
The PRISMA flow diagram of study inclusion/exclusion process.

### Eligibility criteria

The selection of included studies was based on an inclusion/exclusion criteria. Inclusion criteria includes any randomised or observational studies that assessed the concentration level of cytokines and chemokines in peripheral blood of patients with severe SARS, MERS and COVID-19. Severe patients or critical were defined as patients who had a respiratory failure, required mechanical ventilation, shock, combining other organ failures, and intensive care unit (ICU) were needed. Conference proceedings, reviews, studies not published in English, animal studies and *in vitro* cytokine production in stimulated cells were excluded. Studies that did not clearly differentiate between severity or did not report the mean and standard deviation or median of cytokine parameter levels were also excluded.

### Data extraction

Initially, all the researchers (A.Z, A.N, H.A and F.M.) screened the title and abstract of each study independently to assess if it is eligible based on the inclusion/exclusion criteria. Subsequently, all the researchers (A.Z, A.N, H.A and F.M.) conducted a two-step full-text literature search independently to assess eligibility further. Data extraction from the relevant studies was performed independently by all researchers using a form created for this study. The following demographic details were extracted from each study; study details, data source, year of publication, sample size, age and sex. The mean, median, standard error, IQR, and range of the following cytokines and chemokines were also extracted; IL-1, IL-2, IL-4, IL-6, IL-8, IL-10, IL-17, TNF, INFg, RANTS, CCL2/MCP-1, CXCL10/IP10, and TGFß.

### Quality assessment

Methodological quality and risk of bias were assessed using the Newcastle Ottawa Scale for Observational Studies (28), which was modified to meet the requirement of this study (29). A total of six criteria were evaluated: representativeness of the population, sample size, statistical analysis, missing data, methodology to report the outcome of interest and methods to detect or report the outcome of interest. Each criterion was rated using a scale ranging from 0 to 3, where 3 represented the highest quality. The highest possible overall score is 18. In addition, we categorised the quality assessment score into three categories: good quality (>12), moderate quality (>6) and low quality (≤6) points.

### Data Synthesis and Analysis

Descriptive statistics were used to describe the demographic characteristics and the quantitative mean concentration of the inflammatory biomarkers reported in each study. To standardise the data for meta-analysis, we estimated the mean cytokine concentrations from the studies that reported the median using the following formula: calculating the mean: if the sample size 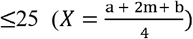, if the sample size >25 (*x* = *m*), and calculating the variance: if the sample size 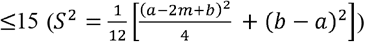, if the sample size 15-70 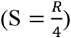 and if the sample size 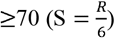(30). A random-effect model was used to estimate the mean concentration of biomarkers (31). Heterogeneity among included studies in the meta-analysis was assessed using the standard χ2 tests and the I^2^ statistic. If high heterogeneity was indicated (I^2^ ≥ 75 %), a systematic narrative synthesis was provided. All analysis was performed using STATA 15.0.

*m = median, a = minimum, b = maximum, X = mean, S = variance, R = range (b-a)

## Results

### Studies selection, characteristics and quality assessment

A total of 4282 studies were identified through the initial online search in databases (Fig 1). Of these, 666 studies were duplicates. Title and abstract screening identified 34 studies for final full-text review using inclusion/exclusion criteria. Finally, the search yielded 18 studies that met the inclusion criteria. Of these, fifteen studies were conducted on COVID-19, two studies on MERS, and one study on SARS. The fifteen COVID-19 studies included 538 severe patients with around (67%) male and (33%) females included in the final analysis. One SARS (47) and two MERS studies (48, 49) had smaller number of patients. The SARS study included 30 severe patients, 70% of them were male and 30 % were females. Moreover, the two MERS studies (48, 49) had 15 severe patients, with round 80% male and 20% females. The final 18 studies also included 1922 non-severe individuals as controls. 13 out of the 18 studies were conducted in China, 3 studies in South Korea, 1 study in Germany, and 1 study in Ireland. Most of the laboratory tests were conducted after admission. The characteristics of the included studies are summarised in Table 1. Overall, the quality of the included studies was good. The quality assessment score for the included studies ranged from 12 as the lowest score to 18 as the highest score recorded. Most of the studies (94.4%) included in this review were of good quality and scored >12 (n = 17), while one study was of moderate quality. The details of the quality assessments are presented in Table 1 and Supplementary table 1

**Table 1.** Characteristics of included studies.

| Author, year of study (ref) | Diseases | Country | Study type | Severe cases |  |  | Non-severe cases |  |  | Quality |
| --- | --- | --- | --- | --- | --- | --- | --- | --- | --- | --- |
|  |  |  |  | Sample size | Age | Study type | Sample size | Age | Male % |  |
| Chen Guang, 2020 (32) | COVID-19 | China | Retrospective study, single centre | 11 | 61[IQR 56.5-66] | 91 % male | 10 | 52.0 [IQR42.8–56.0] | 70% male | Good |
| Gao Yong, 2020 (33) | COVID-19 | China | Retrospective study, single centre | 15 | 45.20±7.68 | 60 % males | 28 | 42.96 ±14.00 | 60% males | Good |
| Han Huan, 2020 (34) | COVID-19 | China | Retrospective study, single centre | 17 | 65.1±14.4 | 52 % males | 42 | 58.3±12.6 | 48% males | Moderate |
| He Susu, 2020 (35) | COVID-19 | China | Retrospective study, single centre | 33 | 54±12.5 | 55 % males | 60 | 44±12.5 | 52% males | Good |
| Herold Tobias, 2020 (36) | COVID-19 | Germany | Prospective study, single centre | 13 | 64 [IQR 45-81] | 91% males | 27 | 58 [IQR18-84] | 58% males | Good |
| Liu Yang, 2020 (37) | COVID-19 | China | Retrospective study, single centre | 30 | NA | NA | 46 | N.A. | NA | Good |
| Luo Miao, 2020 (38) | COVID-19 | China | Retrospective study, two centres | 201 | 69.00[IQR 62.00–78.00] | 66.2 % males | 817 | 57.00 [IQR46.0–66.0] | 47.5% males | Good |
| McElvaney Oliver J, 2020 (39) | COVID-19 | Ireland | Retrospective study, single centre | 20 | 54.3±18.2 | 65 % males | 20 | 56.6±17.3 | 60% males | Good |
| Chen Ruchong, 2020 (40) | COVID-19 | China | Retrospective cohort study, multiple centres | 48 | 61.4±13.6 | 79 % males | 354 | 67.3±12.1 | 52.8% males | Good |
| Wan Suxin, 2020 (41) | COVID-19 | China | Cross-sectional, single centre | 21 | 61.29±15.55 | 52 % males | 102 | 43.05± 13.12 | 53% males | Good |
| Xiaohua Chen, 2020 (42) | COVID-19 | China | Retrospective study, single centre | 17 | 79.6±12.6 | 88.2 % males | 21 | 52.8±14.2 | 61.9% males | Good |
| Yang A. P, 2020 (43) | COVID-19 | China | Retrospective study, single centre | 24 | 57.9±11.8 | 55 % males | 69 | 42.1±18.6 | 75% males | Good |
| Yuan X, 2020 (44) | COVID-19 | China | Cross-sectional, two centres | 46 | 68 [IQR 61-76] | 45.7 % males | 60 | 66 [IQR52-69] | 49.2% males | Good |
| Zhou Yaqing, 2020 (45) | COVID-19 | China | Retrospective study, single centre | 13 | 67.38 ± 13.36 | 77 % males | 8 | 64.00± 15.51 | 37.5% males | Good |
| Zhu Zhe, 2020 (46) | COVID-19 | China | Retrospective study, single centre | 16 | 57.50±11.70 | 56.25 % males | 111 | 49.95± 15.52 | 65.77% males | Good |
| Zhang Yuanchun, 2004 (47) | SARS | China | Retrospective study, single centre | 30 | 45.4 [IQR19–86] | 70 % males | 30 | 44.1 [IQR17–80] | 56.6% males | Good |
| **Hong Ki-Ho, 2018 (48)** | MERS | South Korea | Retrospective study, multiple centres | 6 | 59±8 | 83 % males | 24 | 46±13 | 58% males | Good |
| **Kim Eu Suk, 2016 (49)** | MERS | South Korea | Retrospective study, multiple centres | 9 | 62±13.5 | 77 % males | 8 | 54.25±10.9 | 75% males | Good |

1 The age is expressed as mean ± S.D. or median [IQR].

**Table 2.** Details on the pro-inflammatory and anti-inflammatory cytokines profile among all study participants.

| IL-1 |  | Severe | Non-severe | Significant |
| --- | --- | --- | --- | --- |
| Authors (Ref) | Diseases | ng/ml mean ± SD |  |  |
| Chen Guang (32) | COVID-19 | 5±0.1 | 5±0.1 | No |
| McElvaney Oliver J (39) | COVID-19 | 40.8±10.4 | 13.7±5.8 | Yes |
| Liu Yang (37) | COVID-19 | 6.7±2 | 6.3±1.3 | No |
| Yang A. P (43) | COVID-19 | 38.1±37.4 | 19.5±12.4 | No |
| IL-2 |  | Severe | Non-severe | Significant |
| Authors (Ref) | Diseases | ng/ml mean ± SD |  |  |
| Yuan X (44) | COVID-19 | 3.4±0.41 | 3.5±0.5 | No |
| Han Huan (34) | COVID-19 | 3.4±0.17 | 3.4±0.23 | No |
| He Susu (35) | COVID-19 | 1.1±0.55 | 1.4±1.16 | No |
| Zhu Zhe (46) | COVID-19 | 1±0.22 | 1.04±0.2 | No |
| Yang A. P (43) | COVID-19 | 8.1±6.03 | 2.25±1.01 | Yes |
| IL-6 |  | Severe | Non-severe | Significant |
| Authors (Ref) | Diseases | ng/ml mean ± SD |  |  |
| Yuan X. (44) | COVID-19 | 17.3±5.6 | 9.5±3.1 | Yes |
| Liu Yang (37) | COVID-19 | 37.4±21.8 | 10.7±5.7 | Yes |
| Luo Miao (38) | COVID-19 | 68.7±18.5 | 7.2±2.3 | Yes |
| Chen Guang (32) | COVID-19 | 55.5±27 | 16.6±6.8 | Yes |
| Xiaohua Chen (42) | COVID-19 | 66.4±21.6 | 13.9±6.8 | Yes |
| Gao Yong (33) | COVID-19 | 38.6±10.5 | 12.6±4.8 | Yes |
| Herold Tobias (36) | COVID-19 | 158.7±125.5 | 63.9±52.3 | Yes |
| Chen Ruchong (40) | COVID-19 | 8.7±1.8 | 7.6±0.7 | Yes |
| Han Huan (34) | COVID-19 | 32.3±16.7 | 6.7±0.9 | Yes |
| He Susu (35) | COVID-19 | 12.7±8.2 | 4.6±4.3 | Yes |
| Zhu Zhe (46) | COVID-19 | 26±13.4 | 4.9±1.3 | Yes |
| Wan Suxin (41) | COVID-19 | 37.8±3.9 | 13.4±0.6 | Yes |
| McElvaney Oliver J (39) | COVID-19 | 169.4±35.4 | 45.9±12.4 | Yes |
| Zhou Yaqing (45) | COVID-19 | 17.2±5.6 | 35.3±1.9 | No |
| Yang A. P (43) | COVID-19 | 326.5±299.4 | 30.8±27.4 | Yes |
| Hong Ki-Ho (48) | MERS | 85.3±66.9 | 12.5±13.8 | Yes |
| Kim Eu Suk (49) | MERS | 157±38.3 | 29.5±18.5 | Yes |
| Zhang Yuanchun (47) | SARS | 517±769 | 163±796 | Yes |
| IL-17 |  | Severe | Non-severe | Significant |
| Authors (Ref) | Diseases | ng/ml mean ± SD |  |  |
| Wan Suxin (41) | COVID-19 | 1.16±0.03 | 1.1±0.01 | No |
| Yang A. P (43) | COVID-19 | 3.4±2.5 | 3.8±2.5 | No |
| TNF |  | Severe | Non-severe | Significant |
| Authors (Ref) | Diseases | ng/ml mean ± SD |  |  |
| Chen Guang (32) | COVID-19 | 10.55±0.4 | 7.4±0.8 | Yes |
| Han Huan (34) | COVID-19 | 8.7±2.6 | 5.2±0.4 | Yes |
| He Susu (35) | COVID-19 | 1.13±0.3 | 1.4±0.5 | No |
| Luo Miao (38) | COVID-19 | 11.3±1.7 | 6.9±0.5 | Yes |
| Wan Suxin (41) | COVID-19 | 2.9±0.2 | 4.1±0.5 | No |
| Yuan X. (44) | COVID-19 | 5.1±1.6 | 4.4±0.8 | No |
| Zhu Zhe (46) | COVID-19 | 1.5±0.1 | 1.4±0.1 | No |
| Yang A. P (43) | COVID-19 | 284.2±266.3 | 17.7±13.6 | No |
| Zhang Yuanchun (47) | SARS | 57.8±5.7 | 60.1±4.4 | No |
| INF-γ |  | Severe | Non-severe | Significant |
| Authors (Ref) | Diseases | ng/ml mean ± SD |  |  |
| Han Huan (34) | COVID-19 | 3.4±0.3 | 3.3±0.4 | No |
| He Susu (35) | COVID-19 | 2.1±0.6 | 1.9±0.6 | No |
| Wan Suxin (41) | COVID-19 | 6.9±0.6 | 5.1±0.3 | No |
| Yuan X. (44) | COVID-19 | 2.9±0.4 | 2.9±0.4 | No |
| Zhu Zhe (46) | COVID-19 | 1.9±0.3 | 1.2±0.1 | Yes |
| Yang A. P (43) | COVID-19 | 42.2±37.7 | 19±16.8 | No |
| Zhang Yuanchun (47) | SARS | 86.5±20.4 | 63±20.4 | No |
| IL-4 |  | Severe | Non-severe | Significant |
| Authors (Ref) | Diseases | ng/ml mean ± SD |  |  |
| Yuan X. (44) | COVID-19 | 2.9±0.5 | 2.9±0.44 | No |
| Han Huan (34) | COVID-19 | 3.3±0.2 | 3.4±0.2 | Yes |
| He Susu (35) | COVID-19 | 1.5±0.3 | 1.7±0.4 | No |
| Zhu Zhe (46) | COVID-19 | 1.2±0.4 | 1.9±0.19 | No |
| Wan Suxin (41) | COVID-19 | 1.8±0.1 | 1.7±0.02 | No |
| Yang A. P (43) | COVID-19 | 1.8±0.7 | 2.7±1.7 | No |
| Zhang Yuanchun (47) | SARS | 110±12 | 109±13 | No |
| IL-10 |  | Severe | Non-severe | Significant |

| Authors (Ref) | Diseases | ng/ml mean ± SD | | |
| --- | --- | --- | --- | --- |
| Chen Guang (32) | COVID-19 | 10.8±0.61 | 5.8±1.03 | Yes |
| Han Huan (34) | COVID-19 | 8.7±2.6 | 5.2±0.39 | Yes |
| He Susu (35) | COVID-19 | 4.9±2.6 | 3.5±0.9 | Yes |
| Luo Miao (38) | COVID-19 | 10±1.7 | 5.6±0.3 | Yes |
| McElvaney Oliver J (39) | COVID-19 | 47.3±8.7 | 54.7±7.9 | No |
| Wan Suxin (41) | COVID-19 | 4.6±0.2 | 2.5±0.03 | Yes |
| Liu Yang (37) | COVID-19 | 7.7±1.6 | 5.22±0.22 | No |
| Yuan X. (44) | COVID-19 | 4.8±0.6 | 4.3±0.5 | No |
| Zhu Zhe (46) | COVID-19 | 6.8±1.9 | 3.3±0.4 | Yes |
| Yang A. P (43) | COVID-19 | 12.6±9.6 | 3.9±2.5 | Yes |
| Zhang Yuanchun (47) | SARS | 49.7±12.3 | 47±5.3 | No |

**Table 3.** Details on the chemokines profile among all study participants.

| IL-8 |  | Severe | Non-severe | Significant |
| --- | --- | --- | --- | --- |
| Authors (Ref) | Diseases | ng/ml mean ± SD |  |  |
| Chen Guang (32) | COVID-19 | 34.1±9.02 | 15.8±8.6 | no |
| Luo Miao (38) | COVID-19 | 33.8±6.6 | 12.9±1.8 | Yes |
| McElvaney Oliver J (39) | COVID-19 | 115.5±23.2 | 45.2±12 | Yes |
| Liu Yang (37) | COVID-19 | 43.4±30.2 | 10.2±3.02 | Yes |
| Yang A. P (43) | COVID-19 | 1100.1±994.7 | 131.63±113.6 | Yes |
| Zhang Yuanchun (47) | SARS | 143±41 | 165±51 | Yes |
| CXCL10/IP10 |  | Severe | Non-severe | Significant |
| Authors (Ref) | Diseases | ng/ml mean ± SD |  |  |
| Kim Eu Suk (49) | MERS | 2506.8±876.7 | 327.3±160.4 | Yes |
| Hong Ki-Ho (48) | MERS | 815.5±230.8 | 247.8±35.8 | Yes |
| CCL2/MCP-1 |  | Severe | Non-severe | sig |
| Authors (Ref) | Diseases | ng/ml mean ± SD |  |  |
| Hong Ki-Ho (48) | MERS | 888.8±146.8 | 127.5±26.5 | Yes |

### The level of circulating cytokines in COVID-19, MERS and SARS patients COVID-19

The dynamic changes of circulating cytokines (IL-1, IL-2, IL-6, TNF and IFN-γ, IL-4, IL-10, and IL-17) were analysed in the 18 included studies. IL-6 concentrations were reported in fifteen COVID-19 studies, one SARS study, and two MERS studies. Remarkably, all but one study showed a significant elevation on the level of IL-6 in severe COVID-19 patients compared to non-severe groups. The level of circulating IL-6 was also significantly high in severe SARS patients (517±769 pg/ml) compared to non-severe groups (163±796 pg/ml) (47). Furthermore, the two studies (48, 49) that measured the level of circulating cytokines in severe MERS patients showed an elevation on the level of IL-6 in severe MERS patients compared to non-severe groups.

Other cytokines were substantially elevated in patients with severe COVID-19 patients but not in patients with severe SARS and MERS. For example, three studies (32, 34, 38) out of eight showed a significant elevation on TNF level in severe COVID-19 patients compared to non-severe groups. The concentrations level of IL-10 was also measured in seven studies (33-35, 38, 41, 43, 46), of which ten studies reported a significant difference between severe COVID-19 patients compared to non-severe groups. In contrast, (47) and (48, 49) showed that the level of TNF and IL-10 cytokines were not significantly elevated in patients with severe SARS and MERS compared to a non-severe group, respectively.

Most of the other cytokines were not comparatively high in severe patients with COVID-19, SARS, and MERS. For example, one study (46) out of six showed that the level of IFN-γ was significantly high in patients with severe COVID-19 (1.9±0.3 pg/ml) compared to non-severe groups (1.2±0.1 pg/ml). IFN-γ concentrations were also not elevated in patients with severe SARS than the non-severe group (47). Similar results were seen on IL-4. One study (34) out of six showed that the level of IL-4 in severe COVID-19 patients was significantly higher (3.3±0.2 pg/ml) than non-severe groups (3.4±0.2 pg/ml). However, (47) reported that IL-4 concentration in patients with severe SARS was (110±12 pg/ml), similar to that for a non-severe group (109±13 pg/ml). The level of IFN-γ and IL-4 were not available in any MERS studies.

The concentrations of IL-1, IL-2, and IL-17 were only reported in four (32, 37, 39, 43) COVID-19 studies. Of these, one study (39) showed a significant difference between patients with severe COVID-19 (40.8±10.4 pg/ml) and non-severe groups (13.7±5.8 pg/ml). Similar results were seen in IL-2. One study (43) out of five reported a significant difference between severe COVID-19 patients with an average of 8.1±6.03 pg/ml, and non-severe groups, averaging 2.25±1.01 pg/ml. The only two studies (41, 43) that measured the level of IL-17 in severe COVID-19 patients also showed no significant difference between severe COVID-19 patients and non-severe groups. Altogether, our data suggest that a high level of circulating IL-6, TNF, and IL-10 might be associated with the severity of COVID-19. While increased IL-6 only might be related to the severity of SARS and MERS.

### The level of circulating chemokines in COVID-19, MERS, and SARS patients

The concentrations of circulating inflammatory chemokine IL-8 were reported in five (32, 37-39, 43, 47) COVID-19 studies and one (47) SARS study. Remarkably, four (32, 37-39, 43) out of the five COVID-19 studies reported a significant elevation in the level of IL-8 in severe COVID-19 patients compared to non-severe groups. In contrast, the one study (47) conducted in SARS patients reported a significant reduction in the level of IL-8 in severe SARS patients with an average of 143±41 pg/ml compared to the non-severe group averaging 165±51 pg/ml.

The concentration level of C-X-C motif chemokine 10/interferon-gamma-induced protein 10 (CXCL10/IP10) and chemokine ligand 2/monocyte chemoattractant protein-1 (CCL2/MCP-1) chemokines were only reported in two MERS studies (48, 49). The concentrations level of CXCL10/IP10 and CCL2/MCP-1 were significantly high in patients with severe MERS than in non-severe groups. Although the number of studies measuring the level of chemokines in patients with severe COVID-19, SARS and/or MERS is limited, the data suggest that circulating IL-8 chemokine might be associated with the severity of COVID-19. Moreover, the level of circulating CXCL10/IP10 and CCL2/MCP-1 chemokines might also be related to the severity of MERS.

The 18 studies included in the meta-analysis had a high-level of heterogeneity (I-squared = 99.9 %). There was considerable variation between studies in terms of geographical locations, study designs, time of blood collection, variable assays used to measure the level of cytokines and data source in both severe and non-severe cases (Supplementary figures 1)

## Discussion

Emerging studies focused on selectively targeting elevating inflammatory cytokines for treating cytokine release syndrome in hCoVs (50). However, the mutual cytokine profile among hCoV stains, which could improve the hyperinflammatory state in critically ill hCoV patients is undetermined. In addition to unanswered questions about the mechanistic role of the cytokine storm shared between the three hCoVs. Therefore, this systemic review and meta-analysis were performed to analyse circulatory cytokines and chemokine profiles in patients with severe hCoVs.

Among the inflammatory parameters, our systematic review analysis demonstrated a marked elevation in the level of circulating IL-6 and TNF in severe COVID-19 and MERS patients in comparison to non-severe groups. Most of the included studies also showed a significant elevation of circulating IL-10 in severe COVID-19 patients compared to non-severe groups. However, due to the limited number of studies conducted on SARS, we could not find a strong association of high levels of circulatory IL-6 and TNF and IL-10 with the severity of the disease, which could be an essential factor for further investigation. A previous study on mice showed that SARS spike protein induced the secretion of high levels of IL-6 and TNF through the NF-kB pathway (51). Others indicated that MERS infected human monocyte derived-macrophages induced the secretion of high levels of TNF and IL-6 (52).

IL-6 and TNF are pleiotropic cytokines produced by different cell types during the acute phase of the infection (53, 54). Apart from its pleiotropic property, IL-6 deteriorates inflammation reaction and activates the coagulation pathway by inducing complement components and several acute-phase proteins including, C-reactive protein (CRP), antitrypsin, and fibrinogen (55). Previous studies have also indicated that the high level of circulatory IL-6 is considered an early marker of morbidity and mortality in lung diseases (56). A recent study by Zhua et al. reported the significant role of IL-6 as an independent COVID-19 risk factor using multivariate logistic regression analyses (46). Other studies suggested that the considerable elevation of IL-6 cytokine in severe COVID-19 patients is linked to the massive mucus production by stimulating the expression of the two predominant mucin genes (MUC5AC and MUC5B) in tracheobronchial epithelial cells (57, 58). IL-6 elevation might also be related to endothelial activation and precipitation of a pulmonary immune-mediated thrombosis (59). Moreover, IL-6 has been found to be significantly correlated with the anti-inflammatory cytokine, IL-10, which may reflect self-protection during cytokine release syndrome (46). It has also been reported that the level of anti-inflammatory cytokine could directly contribute to elaborate sepsis-induced immunosuppression, which amplifies the susceptibility to secondary infections (60). Consequently, the intense focus on treating severe COVID-19 cases with an anti-IL-6 receptor (IL-6R) is emerging and showed beneficial results in clinical trials (61).

Less is known about chemokines spectrum in human coronavirus compared to cytokine production. Thus, it is hard to determine the importance of chemokines in hCoVs disease progression. However, our systematic review showed similar or higher levels of chemokines detected in hCoVs infected patients. A marked overproduction of plasma IL-8 was demonstrated in patients infected with severe COVID-19. Whereas CXCL10/IP10 and CCL2/MCP-1 were detected in the blood of patients infected with severe MERS. However, other studies that were not included in our systematic review reported elevated levels of IL-8 in the blood of patients infected with MERS and SARS (52, 62, 63) and CXCL10/IP10 and CCL2/MCP-1 in SARS (64) and COVID-19 (65, 66) infected patients.

IL-8 and CCL2/MCP-1 are secreted by many cells in response to IL-6 and TNF mediated cytokines (67, 68). Whereas CXCL10/IP10 is secreted by many cells, including monocytes, endothelial cells and fibroblast in response to IFN-γ (69). These chemokines and others play essential roles in the pathogenesis of diseases characterized by thrombosis in addition to monocytes, macrophages, neutrophils and lymphocytes infiltration (70). Moreover, anti-CXCL10/IP10 is in a clinical trial as a promoing approach to treat cardiovascular event (71). Thus, the findings of high levels of chemokines in the serum of critically ill hCoVs patients provide some hints that chemokines could be used as biomarkers of the severity of hCoV diseases and can serve as targets for the development of hCoVs chemotherapeutics, especially in patients with thrombotic events.

Our demographic data also demonstrated that most of the hCoVs severe cases are among old male people, which is also in line with previous studies. It has been previously known that older people with comorbidity are more vulnerable than young people to develop a more advance and severe human coronavirus related disease (72). It is believed that dysregulated innate immunity and deficiency in adaptive immune response in older people, mostly males, could play an essential role in this observation (72). For instance, Torcia et al. demonstrated that peripheral blood mononuclear cells (PBMCs) from males stimulated with influenza and Herpes-simplex-1 viruses induced the production of high levels of IL-10 compared to females (73). Other studies showed that males had higher levels of proinflammatory cytokines (e.g., TNF) and chemokines (e.g., CXCL10) following lipopolysaccharide stimulation (74). In contrast, females had a higher number of proliferating T-cells and B-cells *in vitro* and antibody response to influenza vaccination than males (75). Previous studies also demonstrated that patients with comorbidities such as diabetes, asthma, and hypertension are at higher risk of mortality because of the excessive production of inflammatory cytokines, including IL-2R, IL-10, and TNF (76-78). These studies and others highlighted that gender, and age-specific innate and adaptive immunity and comorbidity might affect inflammation reaction of hCoVs patients, immunotherapy, and immune response to hCoVs vaccination.

### Limitation of the study

Several limitations exist within our study; the most important is the observational nature of studies and significant heterogeneity in study results. However, high statistical heterogeneity is more frequent in meta-analyses of prevalence and descriptive studies (79). This can be explained because of the different patient populations, underlying comorbidities and coinfections, variant treatments and follow-up. The different time of blood collections, control donors, and cytokine detection assays can also explain the heterogenicity. Another significant limitation is the variability in laboratory assays used to measure the level of serum cytokines, as local laboratories have different normal ranges based on local data. This confounding variable can somewhat undermine our results and thus, our data should be interpreted as such, keeping in mind this important limitation.

The number of studies included on SARS and MERS compared to COVID-19 were low. However, this can be explained as the COVID-19 infection caused a major threat to the whole population compared to SARS and MERS.

## Conclusions and future directions

Despite these limitations, our study in meta-analysis remained consistent with observational data analysis, demonstrating the importance of cytokine release syndrome and worsen the disease outcome in the three hCoV infectious diseases. Although not all the included studies demonstrated a significant elevation in the level of pro-inflammatory and anti-inflammatory cytokines such as IL-6, IL-10 and TNF, in severe compared to non-severe groups. It is also worth considering that the less noticeable cytokine elevations in the three hCoV infections might suggest a regulated or insufficient inflammatory response to overwhelming viral infection such as IFN-γ.

Although the degree of cytokinaemia in patients with severe SARS, MERS, and COVID-19 is markedly higher than that seen in non-severe groups, there are still unanswered questions regarding the mechanistic role of cytokine storm shared in the three hCoVs. We believe that the immune response and immunopathology in COVID-19 are similar to some extent to SARS and MERS.These findings could shed light on the cytokine release syndrome in the progression of hCoVs and may indicate the type of immune response or pathophysiological events involved in these diseases with a potential therapeutic strategy to improve patients’ outcomes. We recommend using a combination of existing, approved therapies with proven safety profiles such as IL-6 blockade signaling (tocilizumab) for the treatment of hyperinflammation during the severe SARS, MERS, and COVID-19 infections

## Data Availability

No furtehr data is available

## Data Availability

All data sets generated for this study are included in the manuscript/supplementary files.

## Author Contribution

Conception and design FM; Acquisition of the data: AZ, AYN, HA, and FM; Analysis and interpretation of the data: AZ, AYN, HA, and FM. Drafting and revising the article: AZ, AYN, HA, and FM. All authors contributed to the article and approved the submitted version.

## Supplementary materials

The Supplementary Material for this article can be found online at:

