## Supplement for "Circulatory Cytokines and Chemokines Profile in Human Coronaviruses: A systematic review and meta-analysis"

### Supplementary Materials and Data

**Search strategy**

Medline

1. Cytokines.mp. or exp Cytokines/

2. exp Inflammation/ or inflammatory biomarker.mp. or exp Inflammation Mediators/ or exp Biomarkers/

3. exp C-Reactive Protein/ or CRP.mp.

4. Procalcitonin.mp. or exp Procalcitonin/

5. exp Blood Sedimentation/ or ESR.mp.

6. exp Ferritins/ or Serum ferritin.mp.

7. (Cytokine* or inflammation* or inflammatory response*, innate or Biomarker* or biochemical marker* or biologic* marker* or clinical marker* or immune marker* or immunologic marker* or laboratory marker* or serum marker* or surrogate end*point* or surrogate marker* or viral marker* or inflammation* Mediator* or CRP or Procalcitonin or calcitonin 1 or calcitonin precursor* polyprotein or calcitonin related polypeptide alpha or calcitonin-1 or pro-calcitonin or procalcitonin or ESR or Blood Sedimentation or erythrocyte sedimentation or erythrocyte sedimentation rate* or Serum ferritin or Ferritins or basic isoferritin or ferritin* or isoferritin).mp. [mp=title, abstract, original title, name of substance word, subject heading word, floating sub-heading word, keyword heading word, organism supplementary concept word, protocol supplementary concept word, rare disease supplementary concept word, unique identifier, synonyms]

8. 1 or 2 or 3 or 4 or 5 or 6 or 7

9. exp Coronavirus Infections/ or COVID-19.mp.

10. SARS-COV2.mp.

11. (COVID-19 or SARS-COV2 or coronavirus infection* or mers or middle east respiratory syndrome or betacoronavirus).mp. [mp=title, abstract, original title, name of substance word, subject heading word, floating sub-heading word, keyword heading word, organism supplementary concept word, protocol supplementary concept word, rare disease supplementary concept word, unique identifier, synonyms]

12. SARS-COV.mp. or exp SARS Virus/

13. (SARS-COV or SARS or sars associated coronavirus or sars coronavirus or sars related coronavirus or sars virus or sars-associated coronavirus or sars-cov or sars-related coronavirus or severe acute respiratory syndrome virus or severe acute respiratory syndrome related coronavirus or severe acute respiratory syndrome-related coronavirus or urbani sars associated coronavirus or urbani sars-associated coronavirus).mp. [mp=title, abstract, original title, name of substance word, subject heading word, floating sub-heading word, keyword heading word, organism supplementary concept word, protocol supplementary concept word, rare disease supplementary concept word, unique identifier, synonyms]

14. exp Middle East Respiratory Syndrome Coronavirus/ or middle east respiratory virus.mp.

15. (mers virus* or mers-cov or middle east respiratory syndrome coronavirus or middle east respiratory syndrome related coronavirus or middle east respiratory syndrome-related coronavirus).mp. [mp=title, abstract, original title, name of substance word, subject heading word, floating sub-heading word, keyword heading word, organism supplementary concept word, protocol supplementary concept word, rare disease supplementary concept word, unique identifier, synonyms]

16. 9 or 10 or 11 or 12 or 13 or 14 or 15

<u>EMBASE</u>

1. Cytokines.mp. or exp cytokine/

2. exp C reactive protein/ or exp biological marker/ or InflammatORY BIOMARKERS.mp. or exp inflammation/

3. Procalcitonin.mp. or exp procalcitonin/

4. exp erythrocyte sedimentation rate/ or ESR.mp.

5. Ferritins.mp. or exp ferritin/

6. 1 or 2 or 3 or 4 or 5

7. COVID-19.mp. or exp Coronavirinae/

8. SARS-COV2.mp.

9. SARS-COV.mp. or exp SARS coronavirus/

10. middle east respiratory virus.mp. or exp Middle East respiratory syndrome coronavirus/

11. 7 or 8 or 9 or 10

12. (Cytokines or inflammation or inflammatory response, innate or biochemical marker* or biologic* marker* or biomarker* or clinical marker* or immune marker* or immunologic marker* or laboratory marker* or serum marker* or surrogate end*point* or surrogate marker* or viral marker* or Inflammation Mediators or CRP or Procalcitonin or calcitonin 1 or calcitonin precursor polyprotein or

calcitonin related polypeptide alpha or calcitonin-1 or pro-calcitonin or procalcitonin or ESR or Blood Sedimentation or erythrocyte sedimentation or erythrocyte sedimentation rate* or Serum ferritin or Ferritins or basic isoferritin or ferritin* or isoferritin).mp. [mp=title, abstract, heading word, drug trade name, original title, device manufacturer, drug manufacturer, device trade name, keyword, floating subheading word, candidate term word]

13. 6 or 12

14. (COVID-19 or SARS-COV2 or coronavirus infection* or mers or middle east respiratory syndrome or betacoronavirus or SARS-COV or SARS or sars associated coronavirus or sars coronavirus or sars related coronavirus or sars virus or sars-associated coronavirus or sars-cov or sars-related coronavirus or severe acute respiratory syndrome virus or severe acute respiratory syndrome related coronavirus or severe acute respiratory syndrome-related coronavirus or urbani sars associated coronavirus or urbani sars-associated coronavirus or MERS or middle east respiratory virus or mers virus* or mers-cov or middle east respiratory syndrome coronavirus or middle east respiratory syndrome related coronavirus or middle east respiratory syndrome-related coronavirus).mp. [mp=title, abstract, heading word, drug trade name, original title, device manufacturer, drug manufacturer, device trade name, keyword, floating subheading word, candidate term word]

15. 11 or 14

**Supplementary table 1. The quality assessment score of included studies**

| Author, year of study (ref) | Representative population | Sample size | Appropriate statistical analysis | Missing data | Methodology to report the outcome of interest | Methods to detect or report the outcome of interest | Total | Quality |
|---|---|---|---|---|---|---|---|---|
| **Chen Guang, 2020 (32)** | 3 | 1 | 3 | 3 | 3 | 3 | 16 | Good |
| **Gao Yong, 2020 (33)** | 3 | 1 | 3 | 3 | 3 | 3 | 16 | Good |
| **Han Huan, 2020 (34)** | 3 | 1 | 3 | 3 | 1 | 1 | 12 | Moderate |
| **He Susu, 2020 (35)** | 3 | 2 | 3 | 3 | 3 | 3 | 17 | Good |
| **Herold Tobias, 2020 (36)** | 3 | 2 | 3 | 2 | 2 | 2 | 14 | Good |
| **Liu Yang, 2020 (37)** | 3 | 2 | 3 | 3 | 3 | 3 | 17 | Good |
| **Luo Miao, 2020 (38)** | 3 | 3 | 3 | 2 | 3 | 3 | 17 | Good |
| **McElvaney Oliver J, 2020 (39)** | 3 | 1 | 3 | 3 | 3 | 3 | 17 | Good |
| **Chen Ruchong, 2020 (40)** | 3 | 3 | 3 | 3 | 3 | 3 | 18 | Good |
| **Wan Suxin, 2020 (41)** | 3 | 2 | 3 | 3 | 3 | 3 | 17 | Good |
| **Xiaohua Chen, 2020 (42)** | 3 | 1 | 3 | 3 | 3 | 3 | 16 | Good |

| | | | | | | | | |
|---|---|---|---|---|---|---|---|---|
| **Yang A. P, 2020 (43)** | 3 | 2 | 3 | 3 | 3 | 3 | 17 | Good |
| **Yuan X, 2020 (44)** | 3 | 2 | 3 | 3 | 3 | 3 | 17 | Good |
| **Zhou Yaqing, 2020 (45)** | 3 | 1 | 3 | 3 | 3 | 3 | 16 | Good |
| **Zhu Zhe, 2020 (46)** | 3 | 2 | 3 | 3 | 3 | 3 | 17 | Good |
| **Zhang Yuanchun, 2004 (47)** | 3 | 1 | 3 | 3 | 2 | 2 | 14 | Good |
| **Hong Ki-Ho, 2018 (48)** | 3 | 1 | 3 | 3 | 3 | 3 | 16 | Good |
| **Kim Eu Suk, 2016 (49)** | 3 | 1 | 3 | 3 | 2 | 2 | 14 | Good |

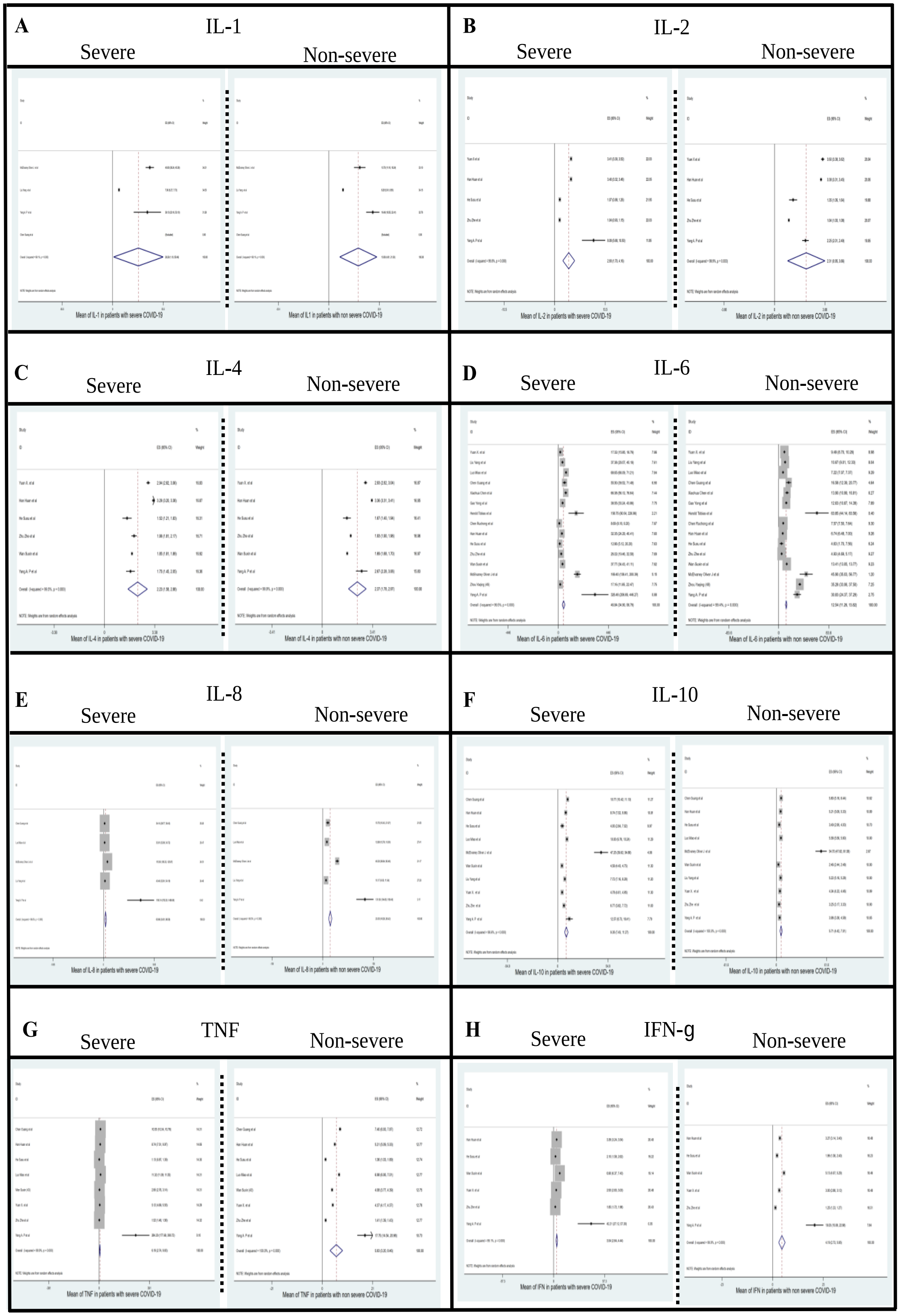


**Supplementary figure 1. A meta-analysis of serum cytokines and chemokines level in COVID-19.** A, IL-1; B, IL-2; C, IL-4; D, IL-6; E, IL-8; F, IL-10; G, TNF; H, IFN-γ.
